# Heart age estimated using explainable advanced electrocardiography

**DOI:** 10.1101/2021.12.08.21267378

**Authors:** Thomas Lindow, Israel Palencia-Lamela, Todd T Schlegel, Martin Ugander

## Abstract

**Background:** Electrocardiographic (ECG) Heart Age conveying cardiovascular risk has been estimated by both Bayesian and artificial intelligence approaches. We hypothesised that explainable measures from the 10-second 12-lead ECG could successfully predict Bayesian 5-min ECG Heart Age.

**Methods:** Advanced analysis was performed on ECGs from healthy subjects and patients with cardiovascular risk or proven heart disease. Regression models were used to predict patients’ Bayesian 5-minute ECG Heart Ages from their standard, resting 10-second 12-lead ECGs. The difference between 5-min and 10-second ECG Heart Ages were analyzed, as were the differences between 10-second ECG Heart Age and the chronological age (the Heart Age Gap).

**Results:** In total, 2,771 subjects were included (n=1682 healthy volunteers, n=305 with cardiovascular risk factors, n=784 with cardiovascular disease). Overall, 10-second Heart Age showed strong agreement with the 5-minute Heart Age (R^2^=0.94, p<0.001, mean±SD bias 0.0±5.1 years). The Heart Age Gap was 0.0±5.7 years in healthy individuals, 7.4±7.3 years in subjects with cardiovascular risk factors (p<0.001), and 14.3±9.2 years in patients with cardiovascular disease (p<0.001).

**Conclusions:** Heart Age can be accurately estimated from a 10-second 12-lead ECG in a transparent and explainable fashion based on known ECG measures, without artificial intelligence techniques. The Heart Age Gap increases markedly with cardiovascular risk and disease.

## Background

Cardiovascular disease is a significant contributor to mortality, with related pathological processes often beginning early and progressing silently for many years^1-3^. Many risk factors for developing cardiovascular disease are lifestyle-related^4^. Fortunately, several are modifiable, and risk can therefore be reduced, for example by smoking cessation, dietary changes and increased physical activity^5,6^. To accomplish this, an individual must fully understand their risk and become motivated to reduce it. One way of communicating risk to the patient is to present it as a “Heart Age”, which can be contrasted to the patient’s chronological age, i.e. a “Heart Age Gap”. A Heart Age can either be determined by translating risk factor scores to what age a resultant score would represent in an individual with no risk factors, or it can be based on electrocardiographic changes^7-12^. Describing the risk to the patient using Heart Age has been reported to reduce metabolic risk factors and may have the advantage of being easily understood by the patient^11,13^. A similar approach has been applied when conveying risk to smokers by describing how ‘old’ their lungs are, and such an approach increased the chance of smoking cessation^14^.

Moving beyond but not excluding basic ECG measurements such as heart rate and waveform amplitudes and durations, the diagnostic output from the ECG can be further substantially improved by also utilizing combinations of advanced ECG measures, for example those from 12-lead-ECG-derived vectorcardiography, waveform complexity, and measures of beat-to-beat waveform variablility^15-17^. In 2014, such advanced measures were used along with 5-minute, high-fidelity 12-lead ECGs and a Bayesian statistical approach, to develop an accurate estimate of ECG-based Heart Age. The estimation of Heart Age using this method has been described in detail elsewhere^9^. In brief, the Bayesian approach was used to infer the deviation of the Heart Age from the subject’s known chronological age by assessing and quantifying an individual’s expected versus actual ECG findings. In addition to sex and chronological age itself, the ECG parameters used in the method as modifiers of the Bayesian-predicted Heart Age were the T-wave axis in the frontal plane, P-wave duration, frontal plane vectorcardiographic QRS axis, spatial JT interval, spatial mean QRS-T angle, high-frequency QRS root mean squared voltages across signal-averaged leads after band-pass filtering, beat-to-beat QT- and RR-interval variability, and measures of T-wave complexity based on singular value decomposition and signal averaging of the T wave. After being trained on a set of healthy individuals, this approach was clinically validated by describing an increased difference between Heart Age and chronological age both for subjects at risk of cardiovascular disease, and those with established cardiovascular disease^9^.

However, importantly, reliable measurement of beat-to-beat RR and QT interval variability requires ECG recordings of at least several minutes. In addition, accurate quantification of high frequency QRS root mean squared voltages requires higher-fidelity recordings with more specialised equipment and sampling rates exceeding the 250-500 samples per second used today in most ECG machines used for standard, 10-second ECG recordings. Therefore, the main aim of this study was to ascertain whether 5-minute ECG Heart Age could be accurately predicted by only those measures available from a standard 10-second 12-lead ECGs. Furthermore, a subsequent secondary aim was to also compare 10-second ECG Heart Ages to chronological ages in healthy subjects, subjects with cardiovascular risk factors, and patients with established cardiovascular disease. We hypothesised that standard-fidelity, 10-second 12-lead ECG recordings could accurately predict Bayesian ECG Heart Ages derived from higher-fidelity, 5-minute 12-lead ECG recordings.

## Methods

A pre-existing database of de-identified subjects with both 5-minute and 10-second 12-lead ECG recordings was utilised for the study^9,16^. Within that database, healthy individuals, patients at cardiovascular risk, and patients with established cardiovascular disease were included. The healthy subjects, who had volunteered as asymptomatic controls, were recruited at Johnson Space Center (USA), the Universidad de los Andes (Venezuela), the University of Ljubljana hospitals and clinics (Slovenia), or Lund University Hospital (Sweden), as previously described in detail^16^. Patients with cardiovascular risk factors or established cardiovascular disease were recruited from cardiology clinics at either Texas Heart Institute (Houston, USA); the University of Texas Medical Branch (Galveston, USA), the University of Texas Health Sciences Center (San Antonio, USA), Brooke Army Medical Center (San Antonio, USA); St. Francis Hospital (Charleston, USA), the Universidad de los Andes (Mérida, Venezuela); and Lund University Hospital (Lund, Sweden).

All healthy subjects were low risk, asymptomatic volunteers with absence of any cardiovascular or systemic disease, based on clinical history and physical examination. Exclusion criteria for the healthy subjects included increased blood pressure at physical examination (≥140/90 mm Hg), treatment for hypertension or diabetes, or active smoking. Patients in the established cardiovascular disease group were included based on the presence of either coronary heart disease (determined by coronary angiography with at least one obstructed vessel (≥50%) in at least one major native coronary vessel or coronary graft, or, if coronary angiography was either unavailable or clinically not indicated, one or more reversible perfusion defects on 99m-Tc-tetrofosmin single-photon emission computed tomography (SPECT)^18-20^), left ventricular hypertrophy (LVH) based on imaging evidence of at least moderate, concentric wall thickening according to guidelines of the American Society of Echocardiography^21^, left ventricular systolic dysfunction (left ventricular ejection fraction ≤50%) at echocardiography, cardiac magnetic resonance imaging (CMR) or SPECT, or findings suggestive of dilated/hypertrophic/ischemic cardiomyopathy at echocardiography or CMR^16^. ECGs were acquired within 30 days of the cardiac imaging examination. Finally, subjects in the cardiovascular risk group were included based on the presence of cardiovascular risk factors such as hypertension or diabetes but no confirmed, established cardiovascular disease^9^.

Based on the above, three groups of study participants were identified: healthy subjects, subjects at cardiovascular risk, and patients with established cardiovascular disease. By methodological design, only healthy subjects were initially included when considering optimal measures of 12-lead ECG available from 10-second recordings for predicting the 5-minute ECG Heart Age. The 10-secondECG measures considered for the prediction model included: (1) From the conventional ECG: heart rate, R-to-R, P-wave, PR, QRS, QT, QTc, and TQ interval durations, as well as the conventional ECG amplitudes and axes; (2) From the transformation of the 12-lead ECG to the Frank X, Y and Z lead vectorcardiogram (VCG) via Kors’ transform^22-26^: the spatial means and peaks QRS-T angles, the spatial ventricular gradient and its individual QRS and T components, the spatial QRS- and T-wave axes(azimuths and elevations), waveform amplitudes and areas, including those in the three individual vectorcardiographic planes, and spatial QRS-and T-wave velocities; and(3) Multiple measures of QRS-and T-wave waveform complexity based on singular value decomposition after signal averaging^27-29^.

For all study participants, the Heart Age Gap was calculated as the 10-second ECG Heart Age minus chronological age. All results were compared between the three groups, and for subgroups based on sex and age (above or below 60 years).

Each participant gave written informed consent. All recordings were obtained under Institutional Review Board (IRB) approvals from NASA’s Johnson Space Center and partner hospitals that fall under IRB exemptions for previously collected and de-identified data. The study was performed in accordance with the Declaration of Helsinki.

### Statistical analysis

Continuous variables were described using mean and standard deviation (SD). The chi-squared test was used to test for proportional differences between groups. Student’s *t* test was used to compare group means.

The 10-second ECG Heart Age was initially derived only in the healthy subject group. First, standard least squares linear regression was used to identify the most promising univariable measures available from 10-second, standard-fidelity ECG for predicting the 5-minute ECG Heart Age. More than 20 such measures were identified, along with chronological age and sex, that had individual P<0.0001. After the univariable linear regression identification procedure, stepwise, standard least squares multiple linear regression was then performed only on the selected measures to best predict, via multivariable model, the 5-minute ECG Heart Age in the healthy subject group. The optimal multivariable model was defined as the most parsimonious one that maintained the highest possible R-squared value while at the same time maintaining all p<0.0001 for the individual measures within the model in the healthy subject group, and at <0.01 when the model was subsequently applied forward to the non-healthy (cardiovascular risk and disease) groups. Comparisons with 5-minuteECG Heart Age are presented as scatter plots and Bland-Altman plots. A two-sided p-value of 0.05 was used as to define statistical significance. Statistical analyses were performed usingSAS JMP version 11.0, SAS Institute Inc, Cary, NC, USA, and R version 3.5.3R, R Foundation for Statistical Computing, Vienna, Austria, https://www.R-project.org/.

## Results

In total, 2,771 patients were included (n=1682 healthy volunteers, n=305 subjects with cardiovascular risk factors, and n=784 with cardiovascular disease). Baseline characteristics are presented in Table 1, and average values for the ultimately included ECG measures are presented in Table 2. The ECG measures included in the final prediction models and the intercept and coefficients for the regression equations are also presented in Table 3 for males and Table 4 for females.

**Table 1.** Baseline characteristics

|  | All<br>(n=2,771) | Healthy<br>(n=1,682) | Subjects at<br>CV risk<br>(n=305) | Patients<br>with CV<br>disease<br>(n=784) | p |
| --- | --- | --- | --- | --- | --- |
| Age, years | 46.2 [16.0] | 38.6 [13.0] | 54.8 [11.2] | 59.0 [13.2] | <0.001 |
| Male sex, n (%) | 1645 [59.4] | 999 [59.4] | 163 [53.4] | 483 [61.6] | 0.048 |
| 5-minute ECG Heart<br>Age, years | 51.1 [21.7] | 38.3 [14.1] | 61.9 [13.2] | 74.2 [15.6] | <0.001 |
| CAD, n (%) | 421 [15.2] | - | - | 421 [53.7] | - |
| ICM, n (%) | 120 [4.3] | - | - | 120 [15.3] | - |
| HCM, n (%) | 92 [3.3] | - | - | 92 [11.7] | - |
| LVH, n (%) | 96 [3.5] | - | - | 96 [12.2] | - |
| NICM, n (%) | 53 [1.9] | - | - | 53 [6.8] | - |

Age and 5-minute ECG Heart Age are presented as mean (standard deviation).
Abbreviations: CAD: coronary artery disease; ICM: ischemic cardiomyopathy; HCM: hypertrophic cardiomyopathy; LVH: left ventricular hypertrophy; NICM: non-ischemic cardiomyopathy

**Table 2.** ECG measures in all patients, stratified by health, cardiovascular risk or cardiovascular disease

| **Measure** | All<br>(n=2,771) | Healthy<br>(n=1,682) | Subjects at<br>CV risk<br>(n=305) | Patients<br>with CV<br>disease<br>(n=784) | p |
| --- | --- | --- | --- | --- | --- |
| P-wave duration, ms | 105 [15] | 99 [13] | 109 [14] | 114 [15] | <0.001 |
| Spatial QT interval, ms | 432 [38] | 428 [32] | 425 [37] | 445 [46] | <0.001 |
| Heart rate, min<sup>-1</sup> | 65 [11] | 63 [10] | 70 [11] | 67 [13] | <0.001 |
| QRS max amplitude in<br>VCG lead Y, μV | 988 [454] | 1144 [392] | 828 [363] | 717 [462] | <0.001 |
| Frontal plane QRS axis,<br>radians | 0.64 [0.44] | 0.82 [0.23] | 0.54 [0.44] | 0.29 [0.54] | <0.001 |
| T-wave complexity, Ln<br>$\sum(\text{EV3:8}) / (\text{EV1}-\text{EV2})$,<br>unitless | -2.9 [0.7] | -3.2 [0.5] | -2.9 [0.6] | -2.3 [0.9] | <0.001 |
| Spatial ventricular<br>gradient minus Spatial<br>mean QRS, mV*s | 0.044<br>[0.030] | 0.059<br>[0.021] | 0.038<br>[0.017] | 0.016<br>[0.028] | <0.001 |
| QRS RMS in VCG<br>vector magnitude lead,<br>mV | 0.52 [0.15] | 0.53 [0.13] | 0.47 [0.11] | 0.50 [0.21] | <0.001 |
| QRS average spatial<br>velocity in VCG vector<br>magnitude lead, mV/s | 45 [12] | 46 [10] | 43 [16] | 43 [10] | <0.001 |
| Portion of QRS loop in<br>posterior superior<br>quadrant of left sagittal<br>plane by VCG, % | 11 [15] | 6 [8] | 11 [15] | 17 [22] | <0.001 |

All values are presented as mean (standard deviation).
Abbreviations: CV: cardiovascular; Ln: Natural logarithm; EV: Eigenvalues; VCG: vectocardiographic; RMS: root mean square

**Table 3.** Measures included in the 10-second ECG Heart Age for male subjects.

| Measure | Estimate | Standard error | t ratio |
| --- | --- | --- | --- |
| *(Intercept)* | -60.575 | 2.353 | -25.73 |
| Age, years | 0.820 | 0.007 | 113.73 |
| P-wave duration, ms | 0.288 | 0.007 | 39.99 |
| Spatial QT interval, ms | 0.081 | 0.004 | 22.86 |
| Heart rate, min<sup>-1</sup> | 0.242 | 0.012 | 20.02 |
| QRS max amplitude in VCG lead Y, μV | -0.007 | 0.000 | -18.20 |
| Frontal plane QRS axis, radians | -5.834 | 0.395 | -14.75 |
| T-wave complexity, $\text{Ln } \sum(\text{EV3:8}) / (\text{EV1}-\text{EV2})$, unitless | 2.152 | 0.164 | 13.12 |
| Spatial ventricular gradient minus Spatial mean QRS, mV*s | -55.841 | 4.464 | -12.51 |
| Male sex (value=1) | 1.996 | 0.215 | 9.27 |
| QRS RMS in VCG vector magnitude lead, mV | 10.315 | 1.612 | 6.40 |
| QRS average spatial velocity in VCG vector magnitude lead, mV/s | 0.106 | 0.017 | 6.30 |
| Portion of QRS loop in posterior superior quadrant of left sagittal plane by VCG, % | 0.033 | 0.009 | 3.55 |
| Abbreviations: Ln: Natural logarithm; EV: Eigenvalues; VCG: vectocardiographic; RMS: root mean square |  |  |  |

**Table 4.**
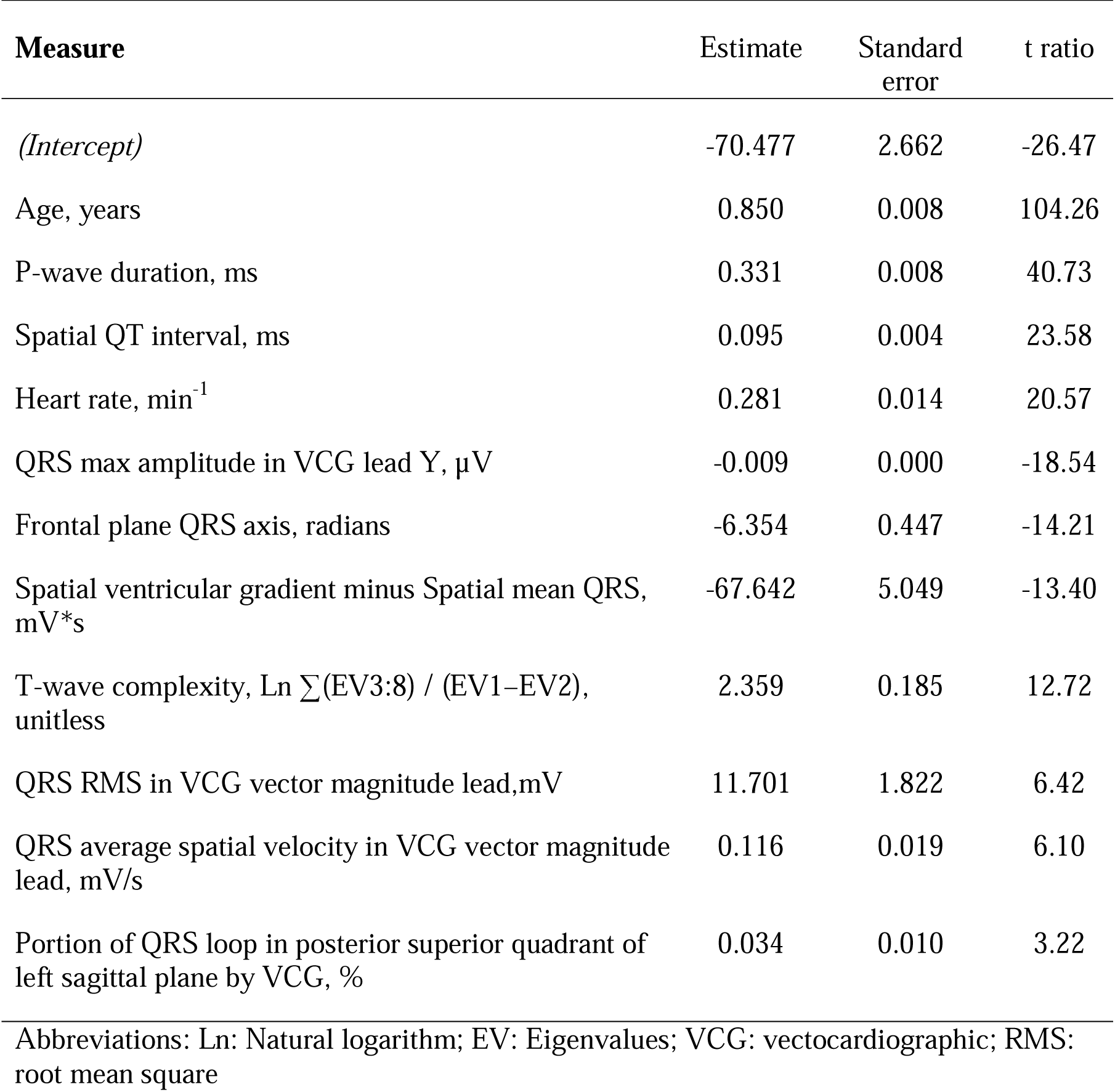
Measures included in the 10-second ECG Heart Age for female subjects.

Agreement between the 10-second Heart Age and the 5-minute Heart Age was excellent (R^2^=0.94, p<0.001, mean±SD bias 0.0±5.2 years), Figure 1. Agreement was strong for both males and females (R^2^=0.91, p<0.001, and R^2^=0.92, p<0.001 respectively). In healthy subjects, on average, the Heart Age and chronological age were no different (0.0±5.7 years), i.e. the Heart Age Gap was zero. In subjects with cardiovascular risk factors, the Heart Age Gap was larger (7.4±7.3 years, p<0.001). Patients with cardiovascular disease showed the largest Heart Age Gap (14.3±9.2 years, p<0.001 vs. subjects at cardiovascular risk), Figure 2. This was evident both for females (Heart Age Gap: healthy subjects: 0.4±5.8 years; cardiovascular risk: 7.8±8.0 years; cardiovascular disease: 14.8±9.0 years, p<0.001) and for males (Heart Age Gap: healthy subjects: -0.3±5.6 years; cardiovascular risk: 7.1±6.6 years; cardiovascular disease: 14.0±9.3 years, p<0.001). A similar pattern was observed for younger (age<60 years) individuals (n=2,142; Heart Age Gap: healthy subjects: 0.1±5.7 years; cardiovascular risk: 7.8±6.4 years; cardiovascular disease: 15.4±9.8 years, p<0.001) and for older individuals (n=629; Heart Age Gap: healthy subjects: -1.7±6.8 years; cardiovascular risk: 7.8±6.4 years; cardiovascular disease: 15.4±9.8 years).

**Figure 1.**
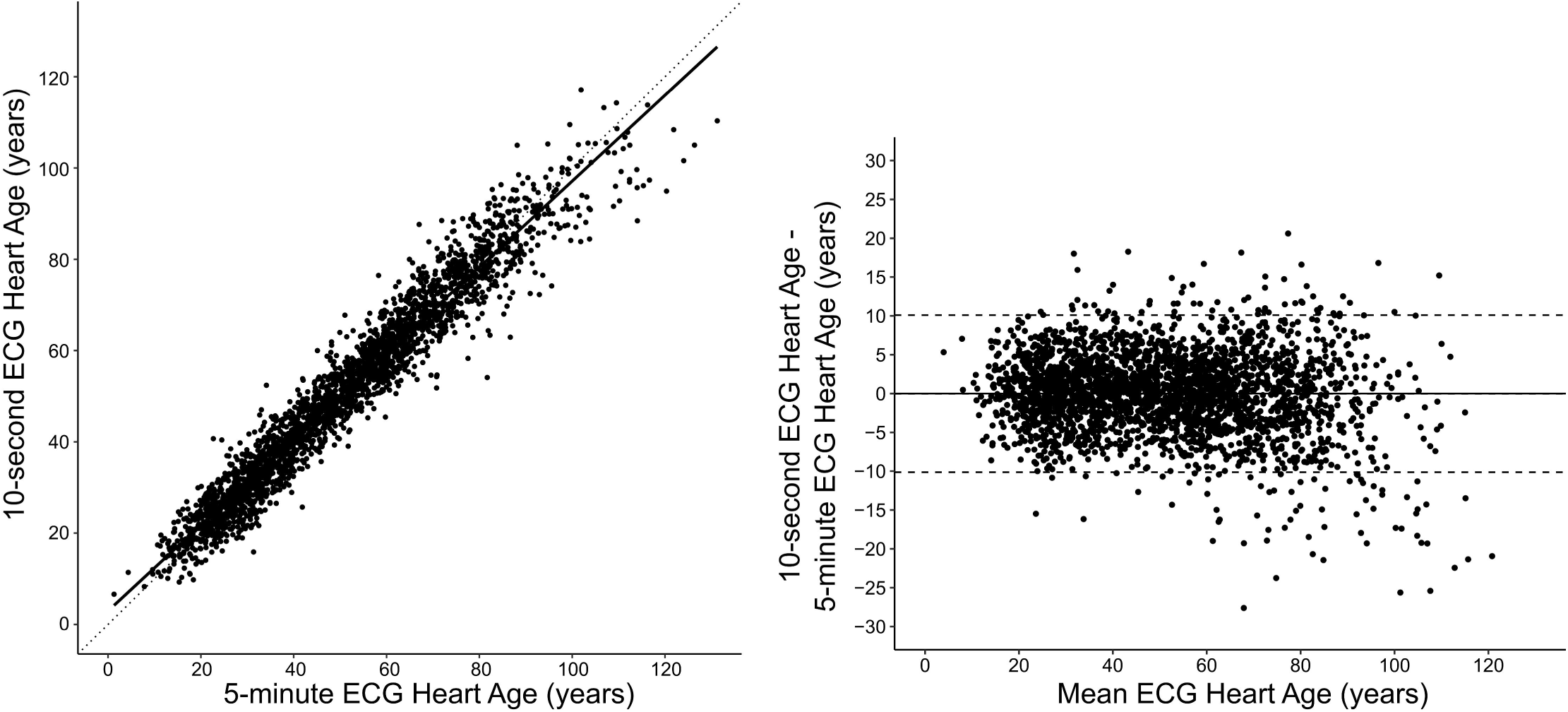
Left panel: Scatter plot showing the relation between the 10-second ECG Heart Age and the 5-minute ECG Heart Age in all participants. The R^2^ value was 0.94 (p<0.001). Right panel: Bland-Altman plot showing the difference between the 10-second and 5-minute ECG Heart Age in relation to the mean of both ECG Heart Ages. The agreement between methods is strong, with minimal deviation from the identity line (dashed) or bias (0.0±5.2 years).

**Figure 2.**
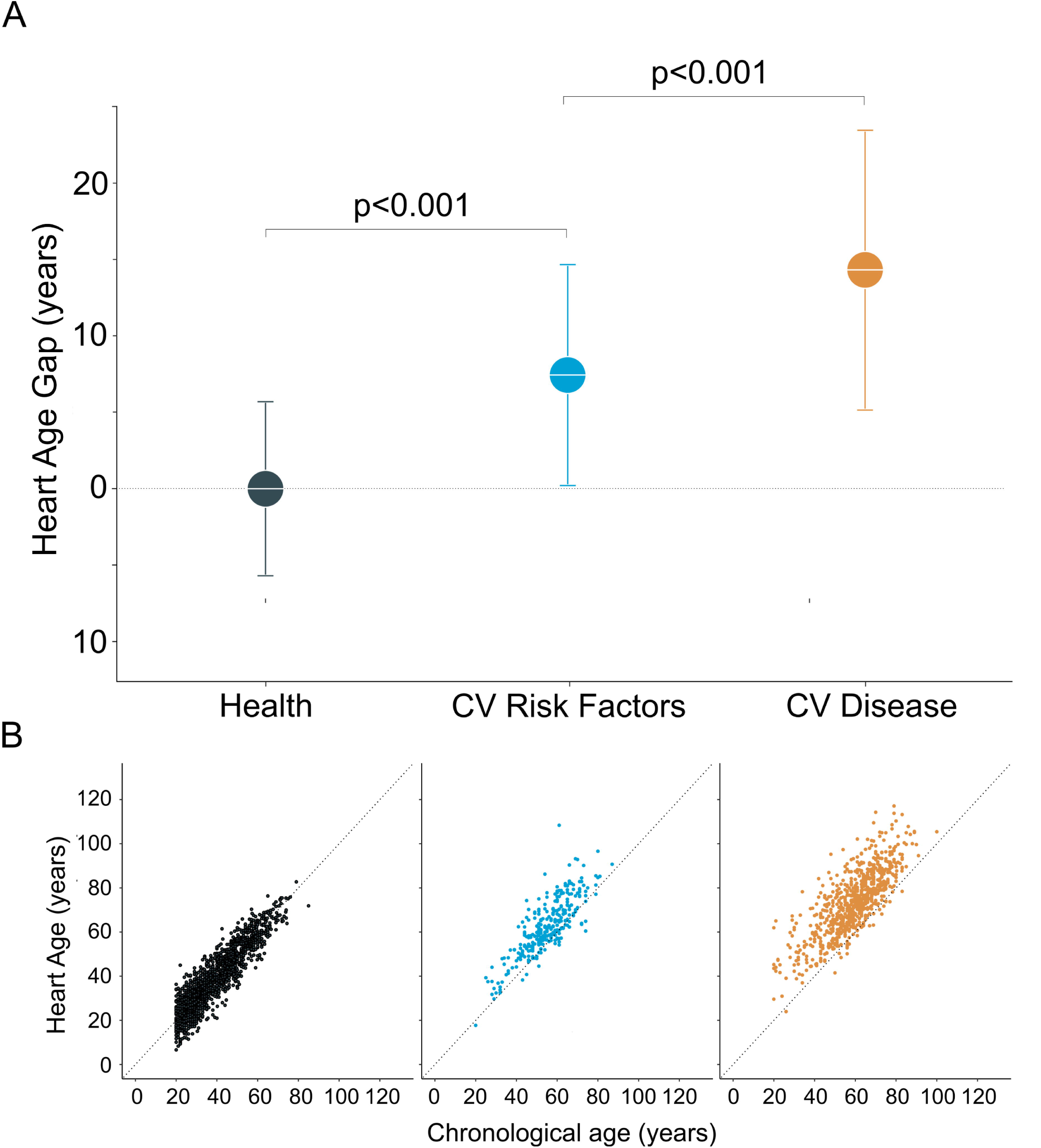
*Panel A:* The difference (the Heart Age Gap) between the 10-second ECG Heart Age and chronological age in healthy subjects (left, dark green), subjects at cardiovascular (CV) risk (middle, light blue), and patients with CV disease (right, yellow).On average, there is no difference between Heart Age and chronological age in healthy subjects. Heart Age Gap is higher in subjects at CV risk, and highest for those with overt CV disease. *Panel B:* Scatter plots showing the relationship between the 10-second ECG Heart Age and chronological age in healthy subjects (left, dark green), subjects at CV risk (middle, light blue), and patients with CV disease (right, yellow). The dashed diagonal line is the identity line, i.e. indicating no difference between Heart Age and chronological age.

## Discussion

We show that Heart Age can be estimated from standard-fidelity, conventional 10-second ECG recordings without requiring more specialised, higher-fidelity, 5-minute long recordings. Consequently, ECG-based Heart Age as estimated using Bayesian techniques can now be more readily implemented in clinical practice.

ECG-based estimation of Heart Age, as a marker of increased cardiovascular risk, has the advantage of being easily understood by the patient. It might also provide strong incentives for lifestyle changes or compliance to medication, similar to the reporting of “lung age” in the context of smoking cessation^14^. For example, in an asymptomatic patient with newly diagnosed hypertension, a value of 160/85 mm Hg may be difficult for the patient to understand in relation to future cardiovascular risk, and the communication of that risk may be difficult for the physician to translate. Instead, if this patient had a chronological age of 50 years but his Heart Age was 10 years higher, it may be more intuitive for the patient to understand that actions are needed. Additionally, an elevated blood pressure is merely a risk factor, and we do not know if the individual patient will suffer from the condition associated with the risk or not. By comparison, changes in the pattern of the ECG and advanced ECG often reflect cardiovascular end-organ changes, some potentially reversible, moving one step beyond the reporting of traditional risk factors.

In addition, we found Heart Age to be similar to chronological age in healthy individuals, while the Heart Age Gap was increasingly larger with increasing cardiovascular disease status. This suggests that Heart Age might also provide useful information for cardiovascular risk prediction, although validation in other datasets is necessary. Moreover, more tailored ECG approaches directly geared toward predicting outcomes, rather than indirectly geared toward predicting outcomes through a Heart Age per se, would also likely outperform strictly Heart Age-based attempts at risk stratification.

In the dataset used, healthy subjects were younger than subjects with cardiovascular risk factors or disease, and there was also a male preponderance in all three groups. The Heart Age Gap showed a similar pattern in both males and females, and in both younger (<60 years) and older subjects.

Estimation of both the 5-minute and 10-second Heart Age currently requires sinus rhythm, since P-wave duration is included in the related calculation. Thus, the assignment of a Heart Age to patients in atrial fibrillation is currently precluded. However, the risk associated with atrial fibrillation is well-established, and patients are often symptomatic. Consequently, the use of Heart Age to incrementally incentivise lifestyle interventions or improve compliance to medications may not be as necessary. Nonetheless, future studies to derive similar methods in patients with atrial fibrillation may also be of value.

### Explainability and transparency of variables that contribute to ECG Heart Age

It is important to note that the patient’s chronological age was purposely included in the estimation of Heart Age, just as in the original (5-min ECG) Bayesian model. The original model was not developed to predict a patient’s actual chronological age, arguably a task of negligible clinical importance. Rather, the model was developed to estimate the “age” of the heart, and specifically how much that estimate differed from chronological age – quantified as the Heart Age Gap. A Bayesian statistical approach is predicated on knowing the patient’s chronological age, with Heart Age being an adjustment of the chronological age based on electrocardiographic characteristics and sex. For an estimation of Heart Age to be accurate in predicting a Heart Age that is similar to the chronological age when the heart is healthy, and increased when the heart is diseased, it is desirable that the included ECG measures change with age, and that the change is augmented with increasing cardiovascular risk or disease severity.

The two ECG measures that had the strongest influence (highest t ratio) on the model were P-wave duration and spatial QT duration. This is not surprising, given that P-wave duration increases with age^30^, and increased P-wave duration can also be seen in advanced cardiovascular pathologies, e.g. heart failure and cardiac amyloidosis^31^. Similarly, QT duration increases with age^32^. Further, QT prolongation is associated with increased of cardiovascular risk, even beyond the rare long QT syndromes^33^, and with incrementally increased risk in advanced ages^34^. These general characteristics are also true for increased heart rate and for leftward shifting of the frontal plane QRS axis^35-38^.The other measures included in the score track changes in the vectorcardiographic QRS and T, and in T-wave complexity by singular value decomposition. Such changes are also known to occur in conditions associated with increased cardiovascular risk, such as hypertension and diabetes^39^, and in established cardiovascular disease, in which they often provide strong diagnostic and prognostic information^22-25,27-29^. Notably, these changes are not easily detectable by visual interpretation of a standard 12-lead ECG. How these ECG measures can affect the Heart Age is exemplified in Figure 3. Taken together, the described ECG measures that contribute in a multivariable fashion to the Heart Age all have physiologically reasonable associations with age and disease in a way that is transparent to the assessing clinician, thus providing important explainability to the model.

**Figure 3.**
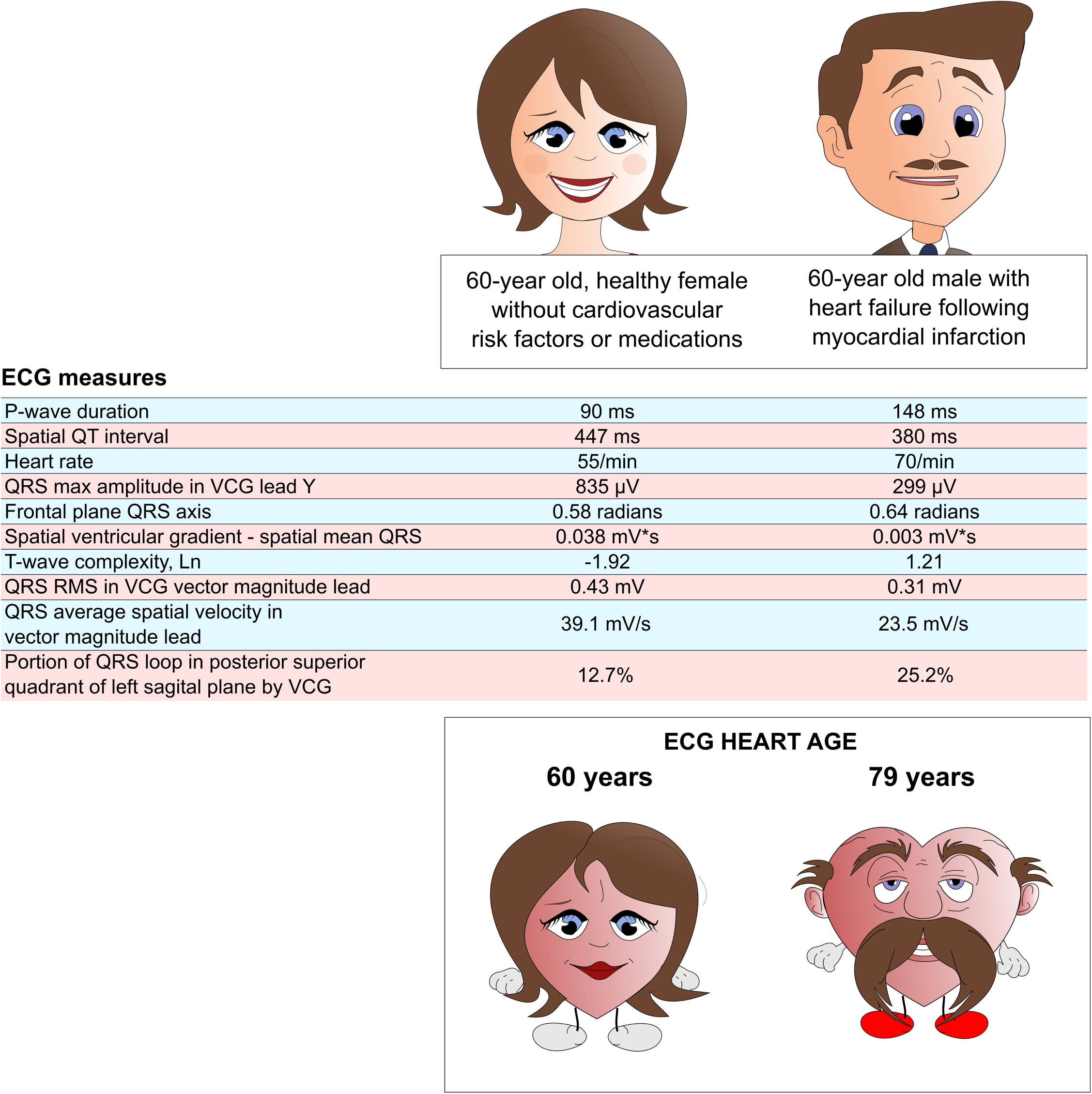
Example of the transparency and explainability of the Heart Age from two subjects with equal chronological age but different Heart Ages, illustrated by a commensurately aged female heart, but a disproportionately aged male heart. The ECG measures for each of the two patients are shown in the table in the middle of the figure, presented in the order of the relative strength (strongest first, based on t ratio [not shown]) of contribution to the Heart Age. Notably, P-wave duration is markedly different between these two patients, and heart rate is higher for the male than the female, helping drive the Heart Age higher in the male. The R-wave amplitude in lead Y is also much larger in the female, and the difference between the spatial ventricular gradient and the spatial mean QRS in the female is also larger, likely due to preserved T-wave amplitudes, contributing to her relatively younger Heart Age. Furthermore, possibly due to ischemic myocardial injuries, T-wave complexity is increased in the male, suggesting that increased myocardial repolarisation heterogeneity also contributes to driving Heart Age higher in the male, in spite of his shorter QT interval.

### Differences compared to Bayesian 5-minute ECG Heart Age

The original, Bayesian 5-minuteECG Heart Age requires information from measures of beat-to-beat heart rate and QT variability,^9^ and of the root-mean square voltage or other aspects of high-frequency (high fidelity) components of the QRS complex^40-42^. However, 10-second-durationrecordingsof standard fidelity do not allow for such measures, and therefore they were not included in the 10-second ECG Heart Age. However, unlike the original Bayesian 5-minute ECG Heart Age, the 10-second ECG Heart Age should be derivable from any standard 12-lead ECG machine, as long as it is sufficiently equipped with software that can quantify the included measures and calculate the 10-second ECG Heart Age. The presented 10-second ECG Heart Age might therefore be anticipated to contribute to more widespread clinical penetration and use.

### Comparison with other Heart (or Vascular) Ages

Different means of expressing a heart or vascular age have recently been published^13^, including the use of information in the 10-second 12-lead ECG to estimate ECG Heart Age based on artificial intelligence (AI)^7,8,10^. For example, Attia, et al, showed that by using a deep neural network (DNN) AI technique, a patient’s chronological age could be predicted, and that if the difference between the predicted and actual age was small, prognosis was good ^8^. When Heart Age by Attia et al’s technique was notably older than the chronological age, the risk of future death was increased^7^. Such results at least superficially correspond to the findings in our study that the Heart Age Gap increased with increasing burden of cardiovascular risk. Another AI method similar to that of Attia et al also more recently reported similarly encouraging results^10^.

Although the results of such AI studies are promising, DNN-based AI techniques are nonetheless inherently problematic in several respects, especially in relation to their lack of transparency and explainability, i.e., the ‘black box’ of AI^43,44^.Without the ability to know the exact features of the 12-lead ECG that are most important in a given DNN model’s output, both interpretability and ethical accountability are compromised^45^. Moreover, it is effectively impossible for a clinician to identify, when critically evaluating the diagnostic output of a DNN-based AI model, the contribution to the result from methodological artifact or bias merely related to noise or to differing technical specifications between different ECG machines^46^. Or the extent to which unanticipated results might merely relate to excess dependency on the particular characteristics of a given DNN AI model’s training set^47^. In addition, a major flaw in all DNN-based AI ECG age models of which we are aware is that their age predictions were made using training datasets that also included individuals with both cardiovascular risk factors and established disease^8,10^. For ECG Heart Age to be used as a marker of potentially reversible cardiovascular disease and risk, it is imperative that ECG Heart Age agrees with chronological age in healthy populations, since it is the deviations from the line of identity in this relationship that form the basis for disease-related assessment and risk.

Hence, we believe that the pursuit of an ECG Heart Age developed from heart-healthy subjects of varying ages, but without a black-box DNN or related AI methodology is valuable, and that the present results provide sufficient confirmation of accuracy to encourage further development. Moreover, that the use of more transparent regression models will also increase the ability of clinicians to better understand the origin of any unexpected result, and to thereafter relay it to the patient with a more convincing sense of trust and accountability^45^. Finally, Heart Age can also now be retrospectively determined for clinical purposes or for retrospective scientific studies whenever raw digital data of acceptable quality are available from stored, standard 10-sec ECG recordings.

### Limitations

The dataset was not strictly divided into a training and test set. However, the fact that the selected ECG measures were derived only in the Healthy group, then applied forward to the other groups wherein the Heart Age Gap was incrementally larger with increasing cardiovascular risk and overt disease, does serve as a form of clinical validation. The prognostic value of the Heart Age Gap was not investigated and requires further evaluation in future studies. The use of three different levels of cardiovascular risk or disease status (healthy, cardiovascular risk without established disease, and established cardiovascular disease) might also be construed as somewhat arbitrary, although they do constitute a logical grouping with incrementally increasing risk from one group to the next. Future studies are nonetheless also required to validate any association between Heart Age and outcomes associated with different cardiovascular pathologies. Finally, although Heart Age was highly accurate, its precision could not be reliably defined in this study. These aspects therefore need to be addressed in future studies.

## Conclusion

We show that Heart Age can be accurately, transparently, and explainably estimated from a standard 10-second, resting 12-lead ECG utilizing multiple, discrete conventional and advanced ECG measures. The Heart Age Gap increases with increasing cardiovascular risk and disease. However, further prospective evaluation in future studies is also required.

## Data Availability

All data utilized in the present study are available upon reasonable request to the authors.

## Notes

**Fundings:** TL is currently under the support of postdoctoral research grants from The Swedish Heart-Lung Foundation (grant no 20200553), the Swedish Cardiac Society, the Royal Swedish Academy of Sciences (grant no LM2019-0013), Women and Health Foundation, Region Kronoberg (grant no 8301), The Swedish Heart and Lung Association (grant no LKH1387), Swedish Association of Clinical Physiology, and the Scandinavian Society of Clinical Physiology&Nuclear Medicine. The study was funded in part by grants (PI Ugander) from New South Wales Health, Heart Research Australia, and the University of Sydney.

### Competing Interest Statement

TTS is owner and founder of Nicollier-Schlegel SARL, which performs ECG interpretation consultancy using software that can quantify the advanced ECG measures used in the current study. TTS and MU are owners and founders of Advanced ECG Systems, a company that is developing commercial applications of advanced ECG technology used in the current study.

### Funding Statement

TL is currently under the support of postdoctoral research grants from The Swedish Heart-Lung Foundation (grant no 20200553), the Swedish Cardiac Society, the Royal Swedish Academy of Sciences (grant no LM2019-0013), Women and Health Foundation, Region Kronoberg (grant no 8301), The Swedish Heart and Lung Association (grant no LKH1387), Swedish Association of Clinical Physiology, and the Scandinavian Society of Clinical Physiology; Nuclear Medicine. The study was funded in part by grants (PI Ugander) from New South Wales Health, Heart Research Australia, and the University of Sydney.

### Author Declarations

All recordings were obtained under Institutional Review Board (IRB) approvals from NASA's Johnson Space Center and partner hospitals that fall under IRB exemptions for previously collected and de-identified data.

